# Baseline immunotypes and immune entropy are indicators of multiple vaccine responsiveness

**DOI:** 10.1101/2024.05.29.24308098

**Authors:** Alper Cevirgel, Marieke van der Heiden, Sudarshan A. Shetty, Markus Viljanen, Martijn Vos, Elske Bijvank, Yannick van Sleen, Celine Imhof, Joeri A.J. Rolwes, Leonard Daniël Samson, Lisa Beckers, Nynke Rots, Josine van Beek, Anne-Marie Buisman, Debbie van Baarle

**Affiliations:** Center for Infectious Disease Control, National Institute for Public Health and the Environment, Bilthoven, the Netherlands; Department of Medical Microbiology and Infection Prevention, Virology and Immunology research group, University Medical Center Groningen, Groningen, the Netherlands; Information Provision Organization, National Institute for Public Health and the Environment, Bilthoven, the Netherlands; Department of Rheumatology and Clinical Immunology, University of Groningen, The Netherlands; Department of Internal Medicine, Division of Nephrology, University of Groningen, University Medical Center Groningen, Groningen, Netherlands

## Abstract

Immune aging is associated with decreased vaccine responses, but biomarkers for vaccine responsiveness remain unidentified. We analyzed immunotypes describing baseline immune cell profiles and their associations with triple vaccine responsiveness to influenza, pneumococcal, and SARS-CoV-2 vaccines in adults aged 25-78 years. Additionally, we developed an innovative measure, immune entropy, to quantify cumulative perturbations in the immune cell subset network. Specific immunotypes associated with either weak or robust triple vaccine responsiveness. In addition, immune entropy was inversely related to vaccine responsiveness regardless of age. In a validation cohort of older adults, higher immune entropy was also associated with a lower antibody response to the BNT162b2 vaccine. A separate cohort of kidney transplant recipients, typically exhibiting diminished vaccine responses, demonstrated significantly increased immune entropy compared to healthy counterparts. Our findings suggest immunotypes and immune entropy as potential indicators to identify individuals at risk for suboptimal vaccine responses, potentially guiding personalized vaccination strategies.

## INTRODUCTION

Aging of the immune system plays a significant role in the prevalence of age-related diseases and co-morbidities observed in older adults ^1,2^. As individuals age, their immune systems undergo a decline in function. This phenomenon, known as immunosenescence, contributes to increased susceptibility to infectious diseases and diminished vaccine responsiveness ^1,3–5^. Identification of individuals with reduced vaccine responsiveness is vital for the strategic implementation of targeted vaccination programs to ensure their protection ^6–8^.

Previous studies aimed to identify individuals at risk for reduced vaccine responsiveness by examining single immune cell subsets, such as Th1, Th17, CD38+ naive B cells, CD38+ memory B cells and MAIT cells ^9–16^. Although some correlations between these immune cell subsets and a specific vaccine response have been reported, ideally, a universal biomarker predicting multi-vaccine responsiveness would be beneficial to target individuals at risk of generally low vaccine responsiveness. However, several reasons complicate the use of single immune cell subsets to identify these low responder individuals. Since vaccine responsiveness is an emergent property of the immune system, a single immune subset does not capture the complexity of the immune network, and it often fails as a robust predictor of vaccine responsiveness ^17–19^. Moreover, aging trajectories differ from person to person and inter-individual immune variation increases with age, resulting in diverse immune phenotypes ^20–23^. This increased heterogeneous state of the immune system in older individuals also challenges the identification of shared immune variables.

Here we employ two innovative measures: (i) immunotypes and (ii) immune entropy, both capturing the total immune subset profile of individuals across a large age range, to discover predictors of low vaccination responsiveness. Immunotypes cluster individuals with similar immune profiles, as previously described ^23^, whereas immune entropy captures the complexity of the immune network and reflects the total perturbations in the immune network in a single variable. Using the unique set-up of the VITAL clinical trials ^23,24^, we aimed to explore and validate the predictive value of both the immunotypes and immune entropy as biomarkers of vaccine responsiveness to multiple vaccines (influenza booster vaccination, primary pneumococcal vaccination, and primary SARS-CoV-2 vaccination) in the same individuals aged 25 to 98 year old ^24,25^. Since immune entropy was introduced as an innovative measure, its associations with immune aging, CMV-seropositivity and sex differences were further investigated.

Our results reveal significant associations between baseline immunotypes and either weak or robust vaccine responsiveness towards multiple vaccines. Intriguingly, also our integrative biomarker immune entropy was associated with vaccine responsiveness overarching multiple vaccines. By introducing and validating comprehensive immune biomarkers that predict humoral immune responses to multiple vaccines, our research sets the stage for identifying individuals susceptible to overall diminished vaccine response. Consequently, these biomarkers derived from the total immune profile hold the potential to develop novel vaccination approaches tailored to individual immune profiles.

## RESULTS

### Study population defined on baseline characteristics

In this study, we included 305 individuals aged between 25 to 98 years who had immune phenotyping data available at baseline and received the Quadrivalent Inactivated Influenza Vaccine (QIV), Prevenar 13 (PCV13) and SARS-CoV-2 vaccines (mRNA-1273 or BNT162b2) from 2019 to 2022 (Figure 1a, Supplementary Figure 1a). Previously participants of the VITAL clinical trials were categorized into nine different immunotypes ^23^. For 173 participants, immunotype categories and pre- and post-vaccination antibody data for triple vaccine analysis (QIV, PCV13, mRNA-1273) were available (Supplementary Figure 1a). These individuals were between 25 to 78 years old, 46% CMV+, and 50% male (Table 1). Participants’ antibody responses to each vaccine were categorized into response quartiles (Q1 to Q4, with Q1 being the lowest and Q4 the highest) based on antibody levels at day 0 and day 28 post-vaccination. We used the maximum residual values adjusted for baseline levels (maxRBA)^26^. Participants were not previously vaccinated with SARS-CoV2 vaccines and only 7 individuals showed Spike-specific S1 IgG concentration above the cut-off level for seropositivity (10 BAU/mL) indicative of previous infection, therefore, instead of maxRBA, day 28 antibody levels were used for antibody response quartiles.

**Figure 1:**
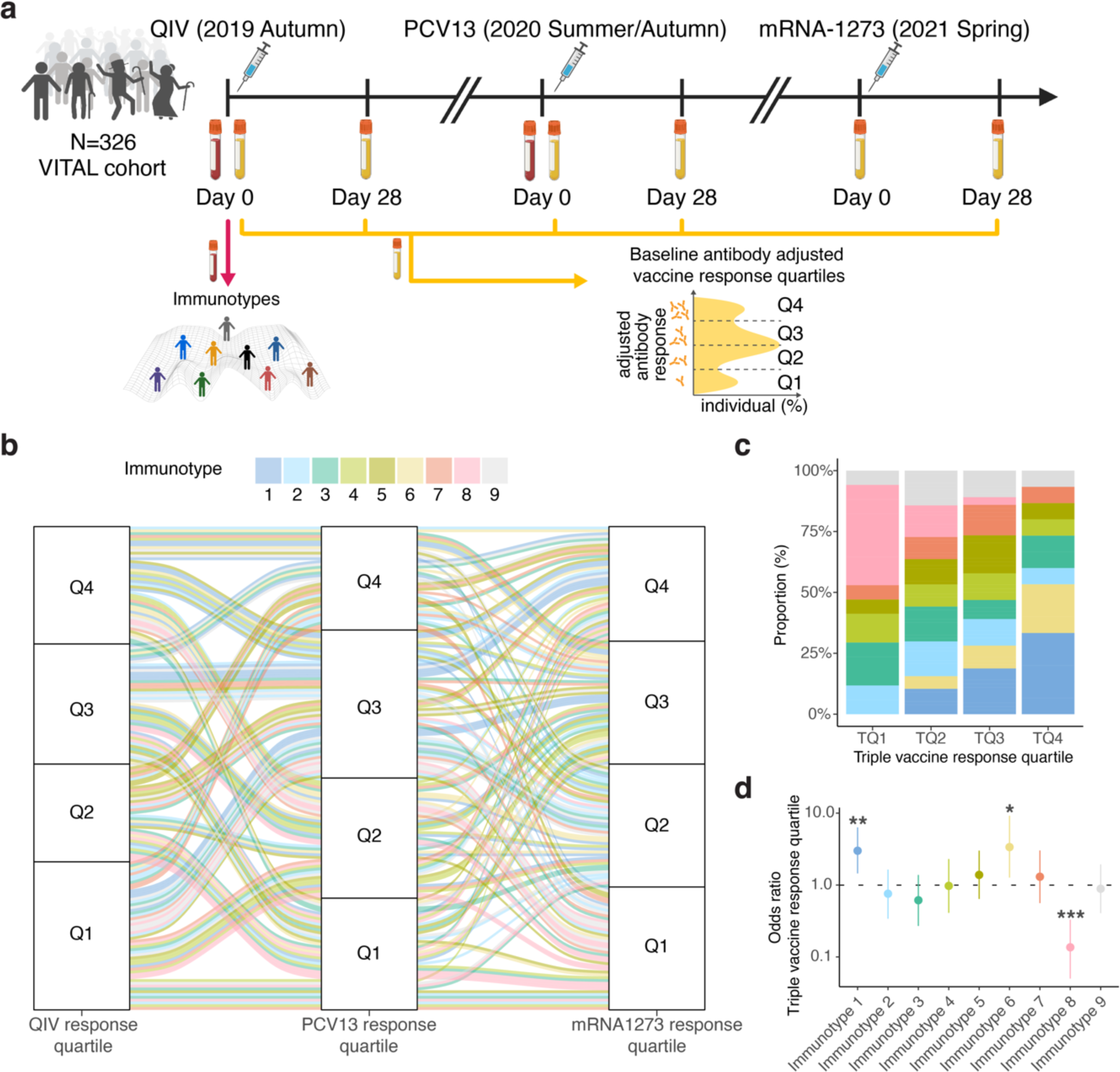
Baseline immunotypes reveal weak and robust antibody responders overarching multiple vaccines. **(a)** Vaccination trial design^24^. Participants’ immunotypes categorized based on pre-vaccination immune subset data^23^. Antibody response groups were categorized in quartiles based on antibody levels for each vaccine at day 28 post-vaccination. Antibody response quartiles indicate lowest (Q1) to highest (Q4) response quartile for each vaccine. **(b)** Antibody response quartiles illustrated across multiple vaccines. Each line represents an individual followed across the vaccines. The line color represents the individual’s immunotype as indicated above. **(c)** Percentages of each immunotype within the calculated triple vaccine response group are displayed. This value is derived from averaging the individual response quartiles across triple vaccines. Triple vaccine response groups (TQ) indicate lowest (TQ1) to highest (TQ4) response quartile. **(d)** Ordinal logistic regression model for triple vaccine response quartiles. \**P* < 0.05, \*\**P* < 0.01, \*\*\**P* < 0.001 showing immunotypes associated with weak and strong vaccine responders.

**Table 1:**
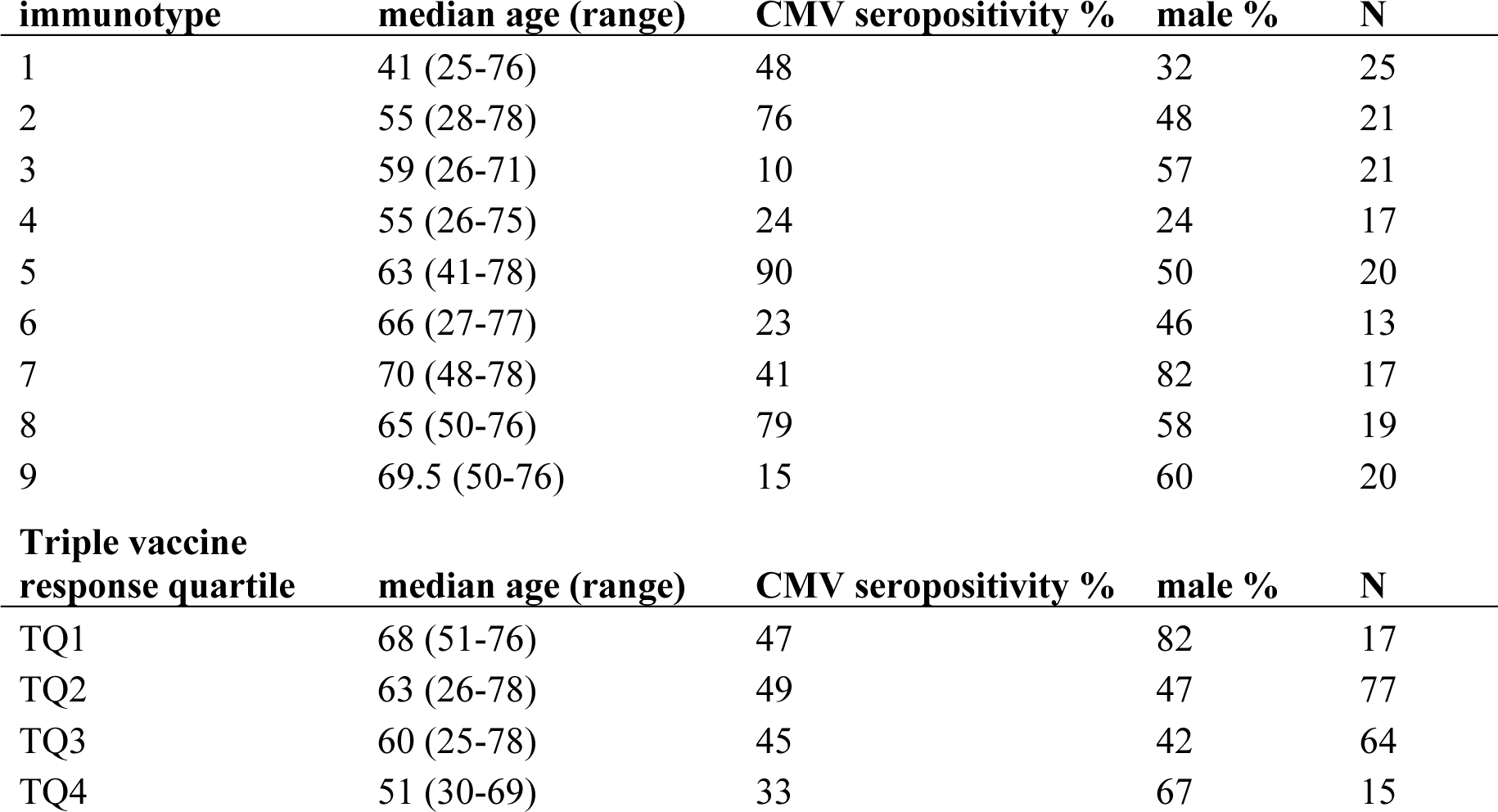
Age, sex and CMV-seropositivity per immunotype in those participants included in the triple-vaccine analysis.

| immunotype | median age (range) | CMV seropositivity % | male % | N |
| --- | --- | --- | --- | --- |
| 1 | 41 (25-76) | 48 | 32 | 25 |
| 2 | 55 (28-78) | 76 | 48 | 21 |
| 3 | 59 (26-71) | 10 | 57 | 21 |
| 4 | 55 (26-75) | 24 | 24 | 17 |
| 5 | 63 (41-78) | 90 | 50 | 20 |
| 6 | 66 (27-77) | 23 | 46 | 13 |
| 7 | 70 (48-78) | 41 | 82 | 17 |
| 8 | 65 (50-76) | 79 | 58 | 19 |
| 9 | 69.5 (50-76) | 15 | 60 | 20 |
| Triple vaccine response quartile | median age (range) | CMV seropositivity % | male % | N |
| TQ1 | 68 (51-76) | 47 | 82 | 17 |
| TQ2 | 63 (26-78) | 49 | 47 | 77 |
| TQ3 | 60 (25-78) | 45 | 42 | 64 |
| TQ4 | 51 (30-69) | 33 | 67 | 15 |

### Baseline immunotypes are associated with vaccine responsiveness overarching multiple vaccines

First, we hypothesized that immunotypes identifying individuals with similar baseline immune profiles are associated with their vaccine responsiveness. Interestingly, we observed a high variability in antibody responses to different vaccines between individuals in all immunotypes (Figure 1b). Only 8% of individuals stayed in the same vaccine response quartile across three vaccines. Meanwhile, 38% differed by one quartile, and 77% differed by two quartiles for at least one vaccine. This variability highlights the importance of studying multiple vaccine responses in the same individual. To this end, we calculated a triple vaccine response quartile (Materials and Methods) and quantified the distribution of the immunotypes in these vaccine response groups. The representation of immunotypes was significantly different for the four triple vaccine response groups (chi-squared p=2.4×10^-2^) (Figure 1c). Interestingly, immunotype 8 was absent in vaccine responders showing the highest antibody response (TQ4), and immunotypes 1 and 6 were not present in those showing the lowest antibody response (TQ1). However, only immunotype 8 showed a significant relationship with the triple vaccine response quartile, comprising 48% of the persons in the lowest response group (Benjamini-Hochberg (BH) p.adj =1.0×10^-4^). We noted a progressive increase in the proportions of individuals with immunotypes 1 and 6 from TQ2 to TQ4. In contrast, the proportions of immunotype 8 were reduced among TQ2 responders compared to those in TQ4. (Figure 1c). These findings imply an association between the pre-vaccination immunotypes and the triple vaccine responsiveness.

To further explore the associations between immunotypes and triple vaccine responsiveness an ordinal logistic regression model was employed. This model revealed that immunotype 1 was associated with increased odds of belonging to a higher triple vaccine response quartile (Confidence Interval (CI)=[1.5-6.3], p=3.4×10^-3^) (Figure 1d). Individuals with this immunotype had a median age of 41, representing the younger adults in our cohort, who lacked aging-related immune perturbations in their immune subset profiles (Table 1)^23^. Furthermore, persons with immunotype 6 also showed significantly increased odds of higher triple vaccine responsiveness (CI=[1.3-9.2], p=1.5×10^-2^) (Figure 1d). Notably, despite their older age (median age of 66) individuals in immunotype 6 resemble more closely individuals in immunotype 1. In addition, these immunotype 6 individuals lack age-related immune subset differences when compared to the mainly older individuals with immunotypes 7, 8 and 9 (Supplementary Figure 2a)^23^. Conversely, persons categorized as immunotype 8 had significantly lower triple vaccine responsiveness (CI=[0.5×10^-1^-0.3], p=1.4×10^-5^). Interestingly, 79% of the individuals with immunotype 8 (median age 65) were CMV-seropositive (Table 1). However, in a separate ordinal logistic regression model, CMV-seropositivity alone was not associated with triple vaccine responsiveness (p=3.9×10^-1^). Although these associations are potentially interesting, immunotypes are categorical variables that describe immune variation in discreet groups, which may not be easily transferrable to other cohorts. Therefore, we aimed to seek a continuous variable that could account for the underlying differences in the immunotypes that are associated with vaccine responsiveness.

### Immune entropy represents cumulative perturbations in the immune cell subset network

Next, we hypothesized that overall immune perturbations in the immune cell subset profile negatively associate with vaccine responsiveness. We propose that the underlying reason why certain persons with immunotype 8 are weak responders is related to a highly perturbated immune network. To capture these immune perturbations in just a single biomarker, we calculated the correlation distance between each individual and a reference group of the youngest individuals within our cohort (<35 years old, N=18) and refer to it as immune entropy. The correlation distance calculation was based on a large set of cellular immune subset data in order to capture the complexity of the immune (Supplementary Table 1). In short, a higher value of immune entropy indicates a greater deviation of the immune profile from that of the reference group and, therefore, a higher degree of immune perturbation. The younger reference group was selected since younger adults typically exhibit a much lower degree of accumulated perturbations in their immune profiles compared to older adults. Within this reference group, the proportions of male and CMV+ individuals were 50% and 28% respectively.

As expected, a significant correlation between age and immune entropy was observed (Spearman’s rho= 0.46, p=2.0×10^-10^, Figure 2a). Moreover, immune entropy was significantly higher in males (p=2.6×10^-3^, Figure 2b), and CMV+ individuals (p=2.9×10^-7^, Figure 2c). Age was not significantly different between males and females or CMV+ or CMV-individuals, and both CMV+ and CMV-individuals showed roughly 50% male/female ratio, suggesting that the differences in immune entropy were not merely driven by the effect of these factors (Supplementary Figure 3a-c). These results indicate that immune entropy reflects the established factors known to influence immune variation. Interestingly, immune entropy was also significantly correlated (BH-adjusted, p.adj<0.05) with serum levels of YKL-40 (Spearman’s rho (r)=0.39, p.adj=4.2×10^-5^), CXCL10 (r=0.31, p.adj=2.5×10^-3^), IL-1RA (r=0.28, p.adj=6.5×10^-3^), CRP (r=0.27, p.adj=6.5×10^-3^), IFNg (r=0.27, p.adj=6.5×10^-3^), IL-6 (r=0.26, p.adj=9.8×10^-3^), Neopterin (r=0.23, p.adj=2.1×10^-2^) and sCD163 (r=0.21, p.adj=4.7×10^-2^) out of an (N=29) inflammatory protein panel known to be associated with low-grade inflammation related to aging. (Supplementary Figure 4a-h). In linear regression models while correcting for age, sex and CMV-seropositivity, immune entropy remained significantly associated with CRP (p=2.2×10^-3^), IL-1RA (p=2.8×10^-2^), TNFa (p=2.9×10^-2^) and CXCL10 (p=4.9×10^-2^). These suggest that immune entropy also reflects age-associated perturbations in serum proteins.

**Figure 2:**
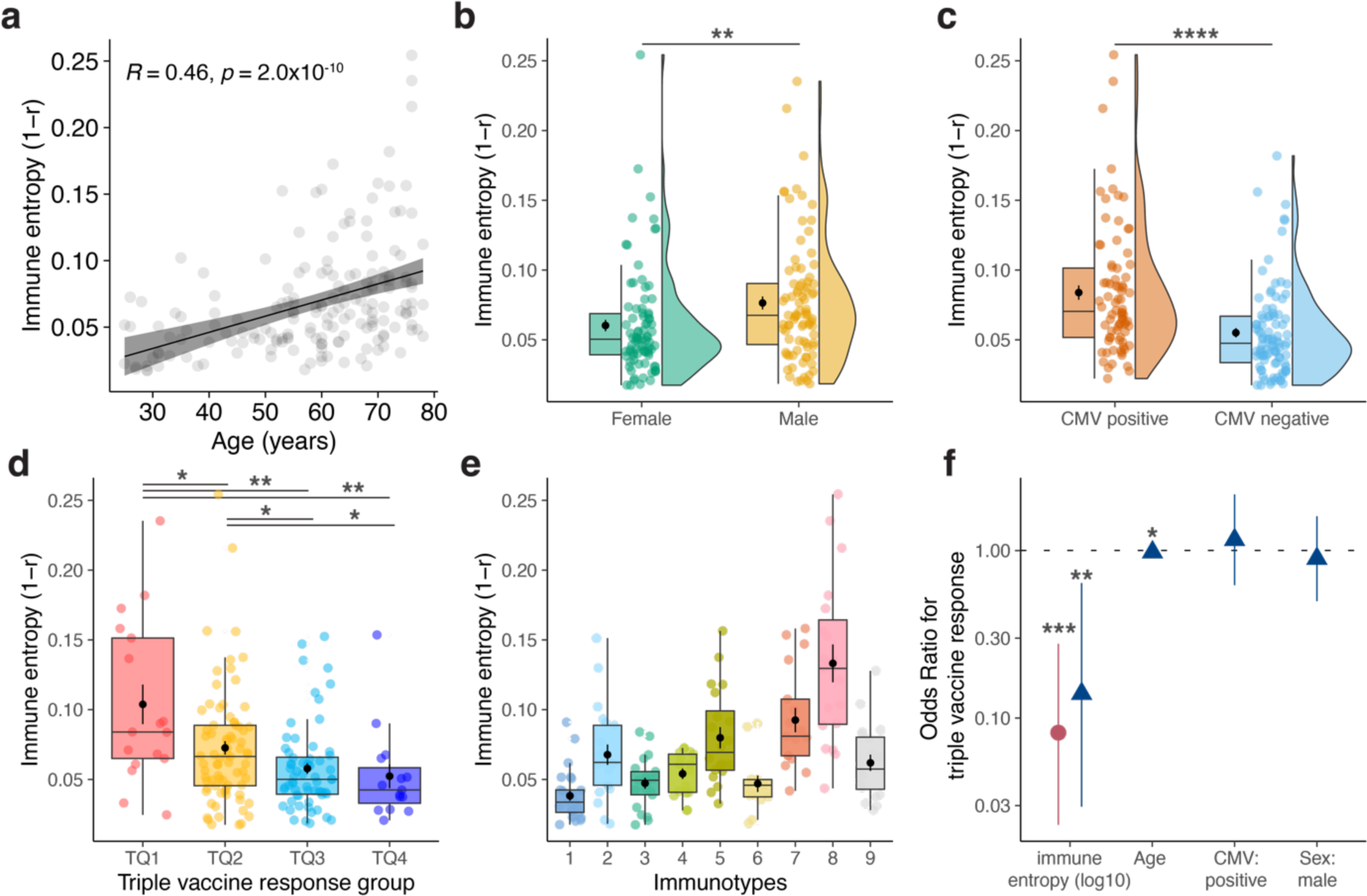
Immune entropy describes the overall immune perturbations and significant associations with triple vaccine responsiveness. **(a)** Spearman correlation between immune entropy and age. **(b)** Immune entropy differences in sex and **(c)** CMV seropositivity. The significance was determined by Mann-Whitney-Wilcoxon test. **(d)** Immune entropy in triple vaccine response quartile groups and **(e)** in immunotypes. The significance was determined using Kruskal–Wallis tests. Post hoc tests were performed using Dunn’s test with the Benjamini-Hochberg method to adjust for multiple comparisons. **(f)** Multivariate ordinal logistic regression model of the associations of immune entropy, age, CMV and sex with the triple vaccine response. The red circle represents the odd ratios for log10 transformed immune entropy, the blue triangle shows the odd ratios for log10 transformed immune entropy, age, CMV seropositivity (CMV positive) and sex (male). \**P* < 0.05, \*\**P* < 0.01, \*\*\**P* < 0.0001,

### Immune entropy is a biomarker of triple vaccine responsiveness

Next, we investigated immune entropy in the context of triple vaccine responsiveness. Immune entropy was significantly higher in individuals classified in the lowest triple vaccine response quartile (TQ1) as compared to all other response quartiles (Figure 2d). Moreover, immune entropy was significantly lower in persons classified into immunotype 8 (weak response associated) as compared to those in immunotypes 1 and 6 (robust response associated) (p.adj<6.9×10^-6^, Figure 2e, Supplementary Table 2). To further investigate the association of immune entropy with vaccine responsiveness, we employed an ordinal logistic regression model. This model revealed that an increase in immune entropy was associated with lower odds of a higher triple vaccine response quartile (CI=[6.8×10^-11^-2.0×10^-4^], p=3.0×10^-5^, Figure 2f red circle). To ensure the robustness of these findings, we conducted a subsequent analysis where we adjusted for potential confounders: age, sex, and CMV-seropositivity in a multivariate ordinal logistic regression model. Even after this correction, immune entropy remained significantly associated with triple vaccine responsiveness (CI=[3.8×10^-10^-1.2×10^-2^], p=3.3×10^-3^, Figure 2f blue triangle). Moreover, in separate ordinal logistic regression models, immune entropy was associated with the response to PCV13 (CI=[2.6×10^-5^-8.6×10^-1^], p=4.4×10^-2^) and mRNA-1273 (CI=[2.8×10^-10^-5.1×10^-4^], p=7.3×10^-5^ separately. This highlights the strength of immune entropy as a biomarker of vaccine responsiveness.

### Immune entropy is associated with BNT162b2 SARS-CoV-2 vaccine response in a validation cohort

To validate the association between immune entropy and vaccine response, we next assessed immune entropy in a different cohort of older individuals who were previously immune phenotyped using a similar panel of markers as used in the VITAL cohort ^27,28^ (Supplementary Figure 1b). A subgroup of these individuals (N=67, aged 64-93) received a primary series of two BNT162b2 SARS-CoV-2 vaccines in 2021 (Supplementary Table 3). Generalized additive models identified a significant non-linear relationship between immune entropy and SARS-CoV-2 Spike-specific antibody levels 28 days after the first dose (p=2.1×10^-3^) and the entire primary vaccination series (p=2.7×10^-2^) (Figure 3a-b). To increase the sample size for this analysis, we added a subgroup of individuals from the VITAL study, also comprising older adults (N=32, aged 72-92), who received the BNT162b2 SARS-CoV-2 vaccine. (Supplementary Figure 1b, Supplementary Table 3). In this consolidated analysis (N=99), immune entropy consistently showed a significant association with Spike-specific antibody levels at day 28 after the first vaccine (p=2.7×10^-2^) and the full primary vaccination series (p=7.9×10^-3^) (Figure 3c, d). These results confirm the significant association of immune entropy with vaccine responsiveness across various cohorts, particularly highlighting its impact on enhancing understanding of the varied responses observed in older adults (aged 65 and above), a group typically characterized by lower vaccine efficacy^29^.

**Figure 3:**
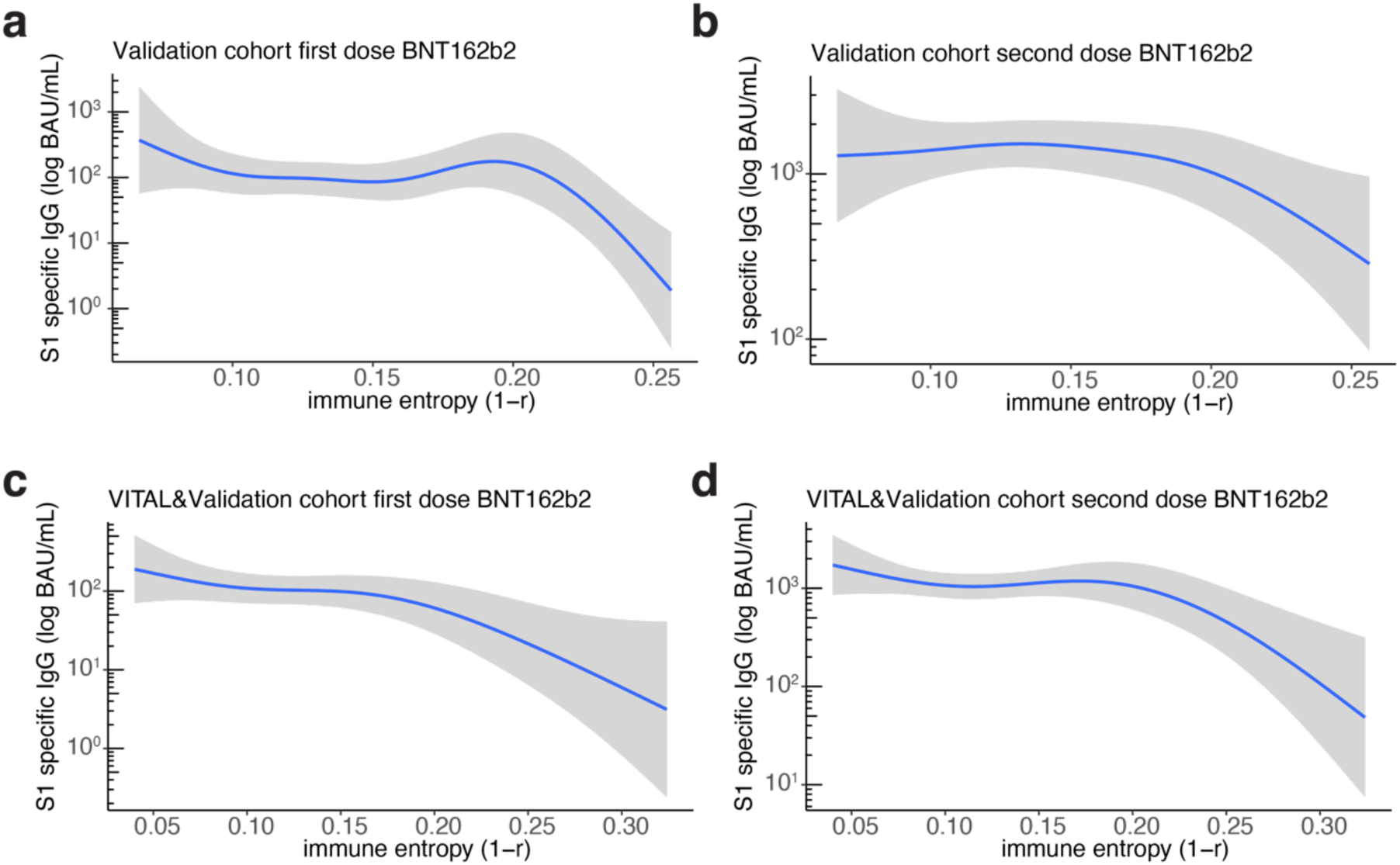
Immune entropy is non-linearly and negatively associated with SARS-CoV-2 vaccine antibody response in a validation cohort of older persons. Generalized additive model of antibody levels to S1 spike protein of SARS-CoV-2 investigating non-linear association with immune entropy after **(a)** first dose and **(b)** the primary series of SARS-CoV-2 BNT162b2 vaccine (N=67) **(c,d)** and after combining BNT162b2 recipients from both the VITAL and the validation cohorts (N=99).

### Kidney transplant recipients show significantly higher immune entropy

Since kidney disease has long been associated with perturbations in the immune system, leading to a heightened vulnerability to infections and a diminished response to vaccinations ^30–32^, we assessed immune entropy in a subgroup of kidney transplant recipients who received a primary series of SARS-CoV-2 mRNA-1273 vaccine (N=59, aged 23-77, Supplementary Table 3, Supplementary Figure 1c) ^33^. We employed a similar immune entropy calculation based on the shared immune cell subset variables between kidney transplant recipients and age-matched generally healthy participants in the present study (VITAL) who received a primary series of SARS-CoV-2 mRNA-1273 vaccine (Supplementary Figure 1a, Supplementary Table 1). Compared to VITAL mRNA-1273 participants (N=194, aged 25-78, Supplementary Figure 1a), immune entropy was substantially higher in kidney transplant recipients (p<2.2×10^-16^, Figure 4a-b). In VITAL mRNA-1273 participants, immune entropy was non-linearly and significantly (p=5.9×10^-7^) associated with Spike-specific antibody levels after the primary series of mRNA-1273 vaccination in a generalized additive model (GAM)(Figure 4c). Here we observed that individuals showing approximately 0.1 and higher immune entropies exhibited a decreased vaccine response, and 97% of kidney transplant recipients who were low responders showing an immune entropy higher than 0.1. These insights extend the utility of immune entropy as a potent biomarker, applicable not just in aging populations but also in disease settings.

**Figure 4:**
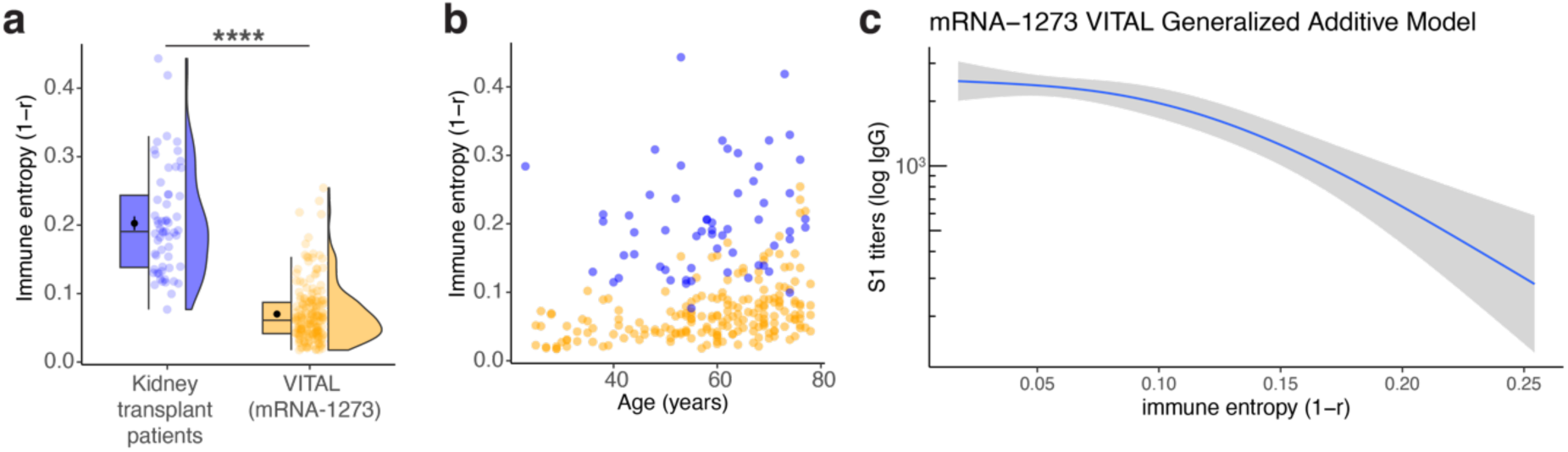
Low SARS-CoV-2 vaccine-responding kidney transplant recipients show increased immune entropy. (**a)** Box and violin plots for immune entropy comparing kidney transplant recipients with generally healthy VITAL participants. The significance was determined by Mann-Whitney-Wilcoxon test. **\*\*\*\****P* < 0.0001.(**b)** Immune entropy and age distribution for kidney transplant patients (blue) and VITAL mRNA1273 (orange) individuals. **(c)** Generalized additive model predictions of SARS-CoV-2 S1 binding antibody units (BAU) 28 days post primary vaccination series and immune entropy in VITAL mRNA-1273 participants.

## DISCUSSION

The heterogeneity in immune aging and the lack of predictive biomarkers of immune function have been obstacles in identifying individuals at risk of diminished vaccine responsiveness and immunosenescence. Our study sought to identify predictors of vaccine responsiveness by analyzing baseline immune phenotypes (immunotypes) and the overall degree of immune perturbations (immune entropy). These variables were derived from an examination of individuals’ total immune cell subset profiles. We showed that specific immunotypes and immune entropy are predictors of responsiveness overarching multiple vaccines, which was independent of sex, age and CMV status

It is postulated that cumulative alterations and remodeling of the immune system over time lead to an accumulation of perturbations in immune profiles, consequently contributing to immunosenescence ^4,34^. While previous research has been valuable in identifying variables associated with aging and immune variation, they fall short in capturing the complexity and integrated nature of immune aging in older adults ^18,35–37^. We propose that conducting a detailed assessment of these immune states that captures both the baseline state and potential responsiveness of the system could be used to better understand immune function and vaccine responsiveness ^38^. We previously categorized baseline immunological states, represented as immunotypes, in our cohort of young, middle aged and older adults ^23^. In this study, we demonstrated that immunotypes 1 and 6 are associated with robust vaccine responsiveness to three different vaccines, in contrast to immunotype 8, which is associated with weak responsiveness. Persons showing immunotypes 1 and 6 represent a mix of both younger and older individuals with strong immune functionality. Notably, characteristics of these immunotypes are a higher naïve-to-memory CD4+ and CD8+ T-cell ratio, and lower HLA-DR+ T cell numbers, as we have previously outlined^23,39^. Key immune characteristics of immunotype 8 included a lower percentage of B cells, a lower naïve-to-memory T cell ratio and higher percentages of follicular (CXCR5+) CD4+ Teff cells, follicular CD4+ Tem cells, follicular CD8+ T cells and follicular CD8+ Tem cells ^23^. Although tissue samples are not available from these individuals to perform further analyses, such striking differences in B cells and follicular T cell compartments could indicate a dysregulation of secondary lymphoid organ structures explaining the observed lower vaccine responsiveness, and these could be potential targets to improve vaccine responses in these people ^40^.

Although immunotypes were associated with vaccine responsiveness, these may not be easily identifiable across different cohorts since the number of immunotypes depends on the initial inter-individual immune variation present in this cohort. Therefore, we sought to determine a continuous measure that could explain the underlying differences between immunotypes in terms of vaccine responsiveness. This measure, termed immune entropy, was associated with factors traditionally linked to immune variation, such as age, sex and CMV-seropositivity^21,23,41^. Previous studies reported that males have lower innate and adaptive immune responses and vaccine responsiveness than females^41–43^. Since males showed significantly higher immune entropy than females irrespective of age and CMV-seropositivity, this could indicate that complex sex-related differences may have an effect on the overall immune perturbations in the immune subset network which were captured by immune entropy. The influence of CMV-seropositivity on the immune cell subset profile is known but its effect on vaccine responsiveness and immune function is still not clearly understood ^22,44^. Since CMV-seropositivity was not associated with triple vaccine responsiveness and the difference in immune entropy could be mainly due to CMV’s effect on the effector T-cell cell compartment, we cannot deduce whether CMV-seropositivity is a confounder or associate of vaccine responsiveness in our analysis.

Immune entropy was negatively associated with responsiveness to PCV13, mRNA-1273 and combined response to QIV, PCV13 and mRNA-1273 (triple vaccine response). This fits with the lack of effect of age on the response to influenza vaccination in comparison to the other vaccines ^24^. Since the VITAL cohort includes adult individuals with a broad age range, immune imprinting, also known as original antigenic sin where immune memory of a pathogen’s initial strain could limit the immune system’s ability to respond to a new strain plays an important role^45^. Moreover, participants in VITAL cohort were vaccinated with the influenza vaccine in previous years. Individuals older than 60 years were vaccinated via regular vaccination campaigns and the younger adults were yearly vaccinated because they were recruited from healthcare workers. Furthermore, recent research highlighted the critical role of memory B cell activation in the efficacy of influenza vaccination^46^. Since our immunotype and immune entropy analysis do not consider these memory B cells in germinal centers, this could explain the lack of association we observed.

Kidney transplant recipients respond poorly to vaccines^33^ and immune entropy was substantially higher in these individuals than in VITAL participants (aged 25-78) suggesting its potential use in clinical scenarios. This elevation in immune entropy may be attributed to the combination of immunomodulatory drugs used post-transplant and the intrinsic immunological challenges presented by kidney disease itself. Immune entropy was also associated with chronic low-grade inflammation markers such as CRP, IL-1RA, TNFa. In cardiovascular disease, chronic low-grade inflammation was suggested to be casually linked to disease progression^47^. Hence, the integration of immune entropy in such disease contexts could provide a more nuanced and comprehensive understanding of patient immune status.

For older adults (aged 65+), previous studies reported diminished and more heterogeneous antibody responses to SARS-CoV-2 vaccines, however, the heterogeneity in these responses is not clearly understood^29^. Interestingly, not only across young to older adults, but also within older adults immune entropy was significantly associated with antibody response to BNT162b2 in the validation cohort (aged 64-93). For these older adults, immune entropy levels higher than 0.2 were associated with a further decrease in BNT162b2 vaccine responsiveness, whereas in the 25 to 78-year-old group who received mRNA-1273, immune entropy levels higher than 0.1 were associated with lower responsiveness. This highlights that immune entropy is applicable when studied in large age ranges to pinpoint individuals at risk for low vaccine responsiveness and also in older adults to gain insights into heterogeneity in antibody response to SARS-CoV-2 vaccines.

Immune entropy is not a single molecule measurement and requires immune subset data obtained via flow cytometry analyses ^23^. Given the widespread use of flow cytometry in clinical contexts, immune entropy could be assessed in various populations and cohorts to explore its associations with vaccine responsiveness and immune function. We propose that future research could help to reduce the number of immune markers required to measure immune entropy, as long as they capture the relevant accumulation of perturbations in the immune subset profile. In our flow cytometry panel, senescent T cells, and various Th populations (Th1, Th2, Th17) were not present. Incorporating such variables that were shown to be important in aging and vaccine responsiveness could further improve the sensitivity of immune entropy and its predictive power ^16,48^.

Although immune entropy was clearly associated with vaccine responsiveness, there were outliers in the higher triple vaccine responder groups that showed increased entropy and vice versa. One reason could be that the set of markers used to calculate immune entropy may not reflect the function of the immune network entirely. Immune entropy calculation using the different collections of immune subsets that would represent the vaccine responsiveness the best could improve this.

Our study underscores the potential utility of both immunotypes and immune entropy as innovative tools to pinpoint individuals at risk of low vaccine effectiveness. This could ultimately guide more personalized vaccination strategies, shifting the healthcare focus from a blanket approach based on age to one rooted in individual effectiveness. This is of high importance to increase vaccine effectiveness as a whole and protect the increasingly vulnerable group of older individuals from infectious diseases.

## MATERIALS AND METHODS

### Study population

This study used samples from the longitudinal intervention studies VITAL and VITAL-Corona as reported in detail elsewhere^24,25^. In short, individuals 25 to 98 years old were recruited. All participants were previously vaccinated with the seasonal influenza vaccination in season 2018-2019 and never received a pneumococcal vaccination. Detailed inclusion and exclusion criteria have been reported^24^. Participants received the seasonal quadrivalent inactivated subunit influenza vaccine in 2019 Autumn (QIV) (2019-2020), and Prevenar 13 (PCV13) in 2020 Summer/Autumn. A group of older individuals received SARS-CoV-2 BNT162b2 (Pfizer) via the national immunization program that started in February 2021 but the majority of participants received the SARS-COV2 mRNA-1273 vaccine (Moderna Biotech, Spain) in 2021 Spring (Supplementary Figure 1a-b) as part of the study via the study team^24^. Ethical approval was obtained through the Medical Research Ethics Committee Utrecht (EudraCT: 2019-000836-24). All participants provided written informed consent and all procedures were performed with Good Clinical Practice and in accordance with the principles of the Declaration of Helsinki. As a validation cohort, participants from the immune system and aging (ISA) sub-study of the Doetinchem Cohort Study (DCS), were used. These individuals were immune phenotyped in 2017 ^27,28^. Part of these individuals (N=67, aged 64-93, Supplementary Table 3) further participated in a SARS-CoV-2 vaccination study (VOCAAL, EudraCT: 2021-002363-22) and received a primary series of SARS-CoV-2 BNT162b2 (Pfizer) vaccinations from February-April 2021. Kidney transplant recipients were part of the RECOVAC study (EudraCT: 2021-000868-30) ^30,33^. A subgroup of these individuals was immune phenotyped (N=59, aged 23-77, Supplementary Table 3) before receiving the primary series of two SARS-CoV-2 mRNA-1273 vaccinations in February-March 2021.

### Serum antibodies and vaccine response profiles

Serum antibody measurements for VITAL and VITAL-Corona studies were described elsewhere in detail^24^. In short, pre and 28 days post-vaccination, antibody levels for each vaccine (QIV: the H3N2 hemagglutination inhibition titer; PCV13: the IgG concentrations against the 13 pneumococcal serotypes; and mRNA-1273: the IgG binding antibody units (BAU) against the Spike S1 protein as measured by Multiplex Immuno Assay were used. Since pre-existing immunity has been shown to influence vaccine response for QIV and PCV13 ^16,39^.vaccine responsiveness was studied rather than day 28 antibody levels,, calculated as the maximum residual values following adjustment for baseline levels (maxRBA) using titer (version 0.0.2) R package^26^. This process adjusts for the initial antibody levels by using a model that describes how antibody levels exponentially increase from their baseline values. Consequently, participants were categorized into vaccine response quartiles (Q1=lowest, Q4=highest) based on their maxRBA. For mRNA-1273, since pre-existing immunity was not present, day 28 antibody levels were directly used to categorize individuals into vaccine response quartiles. A triple vaccine response quartile was determined by averaging the response quartiles for QIV, PCV13, and mRNA-1273. This average was then rounded to define four triple vaccine response categories: TQ1 (lowest), TQ2, TQ3, and TQ4 (highest).

For the validation cohort (ISA), SARS-CoV-2 Spike S1 protein IgG units were measured 28 days for both the first vaccine and the primary series of two vaccinations with respectively Bnt162b2 and with mRNA-1273. The same multiplex platform was used to measure all SARS-CoV-2 Spike S1 protein antibodies in this study ^49^.

### Immune phenotyping and immunotypes

The immune phenotyping data based on a flow cytometry panel of 18 markers utilized in this study has been described in detail in a previous publication ^23^. Briefly, baseline immune subset data (n=59) were subjected to a Spearman correlation matrix analysis to elucidate inter-individual relationships. Subsequent clustering was performed, informed by gap statistics analysis to ensure the optimal number of clusters. The entire methodology can be found in the corresponding GitHub repository (https://github.com/alpercevirgel/Immunotype-Alper2022)^23^. For the validation cohort (ISA), immune phenotyping was performed as described previously ^27^. PBMCs from kidney transplant recipients participating in the RECOVAC cohort were stained using the following anti-human fluorochrome-conjugated antibodies: CD3 (Sparkblue 550, SK7, Biolegend), CD4 (cFluor-YG584, SK3, Cytek), CD8 (BUV805, SK1, BD), CD45RA (Spark NIR 685, HI100, Biolegend), CD95 (BB700, DX2, BD), HLA-DR (BV570, L243, Biolegend), CD38 (APC-Fire810, HIT2, Biolegend), CD19 (eFluor450, HIB19, ThermoFisher), CD27 (VioBright FITC, M-T271, Miltenyi), IgD (BUV395, IA6-2, BD), CD127 (APC-R700, HIL-7R-M21, BD), CD25 (PE-AF700, CD25-3G10,ThermoFisher), CXCR5 (BUV563, RF8B2, BD), CCR7 (BUV616, 2-L1-A, BD), CD28 (BV421, CD28.2, BD) and viability dye (FVS780, BD). Samples were analyzed in Cytek Aurora 5L (Cytek Biosciences) and unmixed using SpectroFlo (v3.1.0). Immune subsets that are used in the immune entropy calculation were exported from FlowJo (version 10.0.7).

### Cytomegalovirus seropositivity

Immunoglobulin G antibodies against CMV were quantified in serum collected before vaccination by a multiplex immunoassay developed in-house^50^. Seropositivity thresholds were adapted from a previous study^44^. For CMV, a concentration of <7.5 relative units (RU) ml^−1^ was categorized as seronegative, ≥7.5 RU ml^−1^ as seropositive.

### Immune entropy

Immune entropy is defined in this study as the correlation distance from a reference group. This distance is calculated as 1 minus Spearman’s rho (Equation 1), indicating the degree of variation in the immune systems of participants when compared to the reference group.

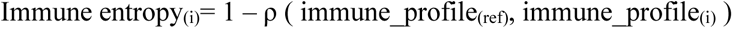

**Equation 1:** Data frame ‘immune_profile’ contains 59 immune cell subset variables (Supplementary Table 1) for each individual, and immune_profile_(i)_ is the vector of 59 variables for a given individual i. ‘immune_profile_(ref)_’ contains the median of all vectors where age_(i)_ <35. ‘ρ’ represents the Spearman correlation coefficient between immune_profile_(ref)_ and immune_profile_(i)_.

For the validation cohort (ISA) and kidney transplant recipients (RECOVAC), immune entropy is calculated using a similar approach. However, due to minor differences in the flow cytometry panels, slightly different immune subset variables are utilized (Supplementary Table 1).

### Serum/plasma protein measurements

From the baseline visit study time point, Angiopoietin-2, C5a, CCL2, sCD25, sCD163, CXCL10, sGP130, IL-1RA, sIL-6R, PTX-3, YKL-40, CRP, sCD14, IL-8, Calprotectin, SAA, Neopterini, FABP2. GM-CSF, IFN-a, IL-1b, TNF-a, IFN-y, IL-6, and IL-10 were measured from serum, and Elastase, PR3, Cathepsin G, and A1AT-Elastase were measured from plasma as described previously ^51^.

### Statistical analysis

Data handling, statistical analyses, and visualization were performed in R (version 4.2.2) and R Studio (version 2022.12.0.353). An ordinal logistic regression was performed to model the triple vaccine response quartile, using immunotypes as an independent variable, and subsequently in a multivariate separate model incorporating immune entropy, age, sex, CMV-seropositivity. For ordinal logistic regression models clm() function from ordinal (version 2022.11-16) package was used. For immunotypes sum coding was used. This method calculates the deviation of each category from the overall mean of categories and is particularly useful when assessing differences from the mean rather than a specific category^52^. Sum coding was implemented by using stats (version 4.2.2) package. Mann-Whitney-Wilcoxon test was used to compare age or immune entropy between sex and CMV-seropositivity differences. The Kruskal-Wallis test from rstatix (version 0.7.2) package was utilized to compare immune entropy across immunotypes. Kruskal-Wallis test was subsequently followed by Dunn’s test when p-values were lower than 0.05. Benjamini-Hochberg false discovery rate correction is applied p-values are reported as p.adj. The correlation between serum proteins and immune entropy was assessed using Spearman’s rank correlation using stats package. Generalized additive models (GAM) were used to study the association between antibody levels to both SARS-CoV-2 mRNA-1273 and BNT162b2 vaccines, and immune entropy. Antibody response was log10 transformed to ensure normality in GAMs. To accommodate potential non-linear associations, immune entropy was modeled as a smooth term. The optimal complexity of the smooth term was determined by selecting the spline’s degrees of freedom (k) that resulted in the lowest Akaike Information Criterion (AIC), with k values ranging from 3 to 20 tested. GAMs were conducted using the mgcv (version 1.9-0) package. In the figures, boxplots display the interquartile range (IQR, 25–75%), with the median indicated by a horizontal line within each box. Whiskers represent the upper- and lower-quartile ±1.5 × IQR. Mean is represented as a black dot and error bars around the mean show the standard error of the mean. These elements combine to provide a detailed visualization of the immune entropy across different immunotypes." Levels of statistical significance are indicated as: \**P* < 0.05, \*\**P* < 0.01, \*\*\**P* < 0.001, and \*\*\*\**P* < 0.0001. Illustrations in the figures are created by using Adobe Illustrator and Biorender.

## Data Availability

The datasets generated during the current studies are not made available in a public database since the study is ongoing. We will share pseudonymized data upon reasonable request as long as data transfer is in agreement with the clinical protocol and EU legislation on the General Data Protection Regulation, which should be regulated in a data sharing agreement.

## Acknowledgment

We would like to thank all collaborators; Inge Pronk, Linde Woudstra, Jacqueline Zonneveld, Sandra Hoogkamer, Marjolein Izeboud, Olga de Bruin, Helma Lith, Joyce Geeber, Ilse Akkerman, Megan Barnes, Lysanne Bakker, Shirley Man, Silvia Cohen, Ruben Wiegmans, Nazela Mir Leibady, Ronith van de Wiele, Rydianne Carvalho De Moura, Emma van Wijlen, Petra Molenaar, Madelène Paets, Martien Poelen, Martijn Vos, Jeroen Hoeboer, Marion Hendriks, Noortje Smits, Jolanda Kool, Maarten Emmelot, Ronald Jacobi, Stefanie Lenz, Floor Peters, Gaby Smits, Marjan Kuijer, Irina Tcherniaeva, Lia de Rond, Mary-lène de Zeeuw-Brouwer, Thierry Ollinger, Wivinne Burny, Rob S van Binnendijk and Marianne A van Houten.

## Author Contributions

NR, JvB, AMB, and DvB conceptualized the study and were involved in funding acquisition.

EB was responsible for data management.

AC, MV, LB, LS, MvdH, YvS, JR were involved in data acquisition.

AC, SAS, MvdH, CI, JR performed data analyses.

AC visualized the data.

AC wrote the original draft.

AC, MvdH, AMB and DvB are involved in writing and editing.

All authors critically revised the manuscript before publication.

## Data and code availability

The codes used in the manuscript are available from GitHub (https://github.com/alpercevirgel/Trivac_entropy)

