## Supplementary Figures and Tables for "Baseline immunotypes and immune entropy are indicators of multiple vaccine responsiveness"

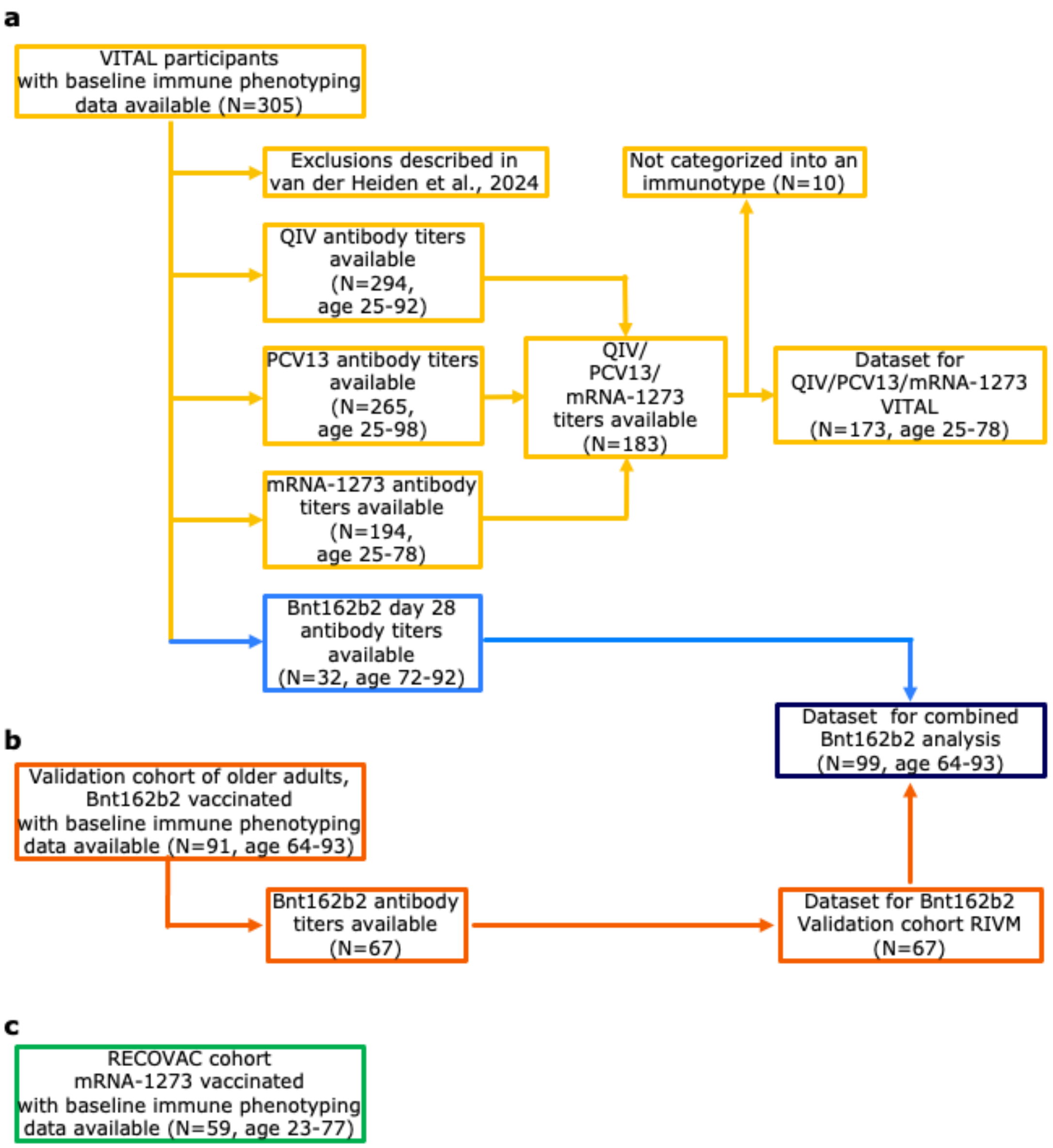


**Supplementary Figure 1: Flowchart of analyses. (a)** Participant flowchart for the VITAL vaccination trial design. Yellow boxes represent participants in the triple vaccine analysis and blue boxes represent Bnt162b2 vaccine analysis. **(b)** Participant flow for the validation cohort Bnt162b2 analysis. **(c)** Participants in RECOVAC kidney transplant recipients.

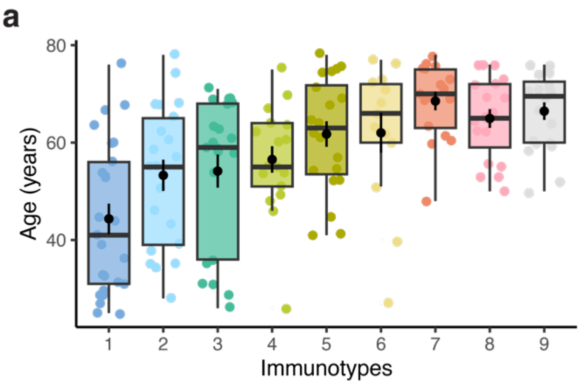

**Supplementary Figure 2: (a)** Age distribution of participants who are included in the triple vaccine analysis per immunotype.

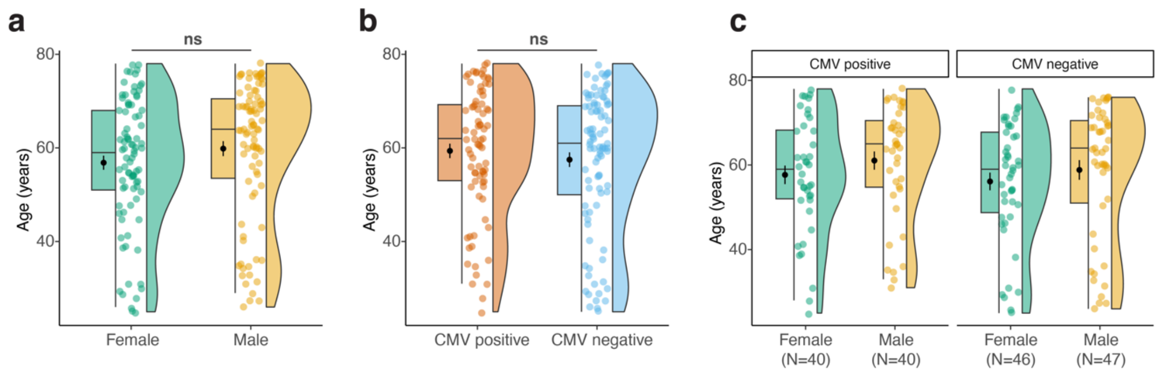


**Supplementary Figure 3: (a)** Age differences between sex, **(b)** CMV-seropositivity, **(c)** between CMV+ males and females and CMV- males and females. The significance was determined by Mann-Whitney-Wilcoxon test. ns $P > 0.05$

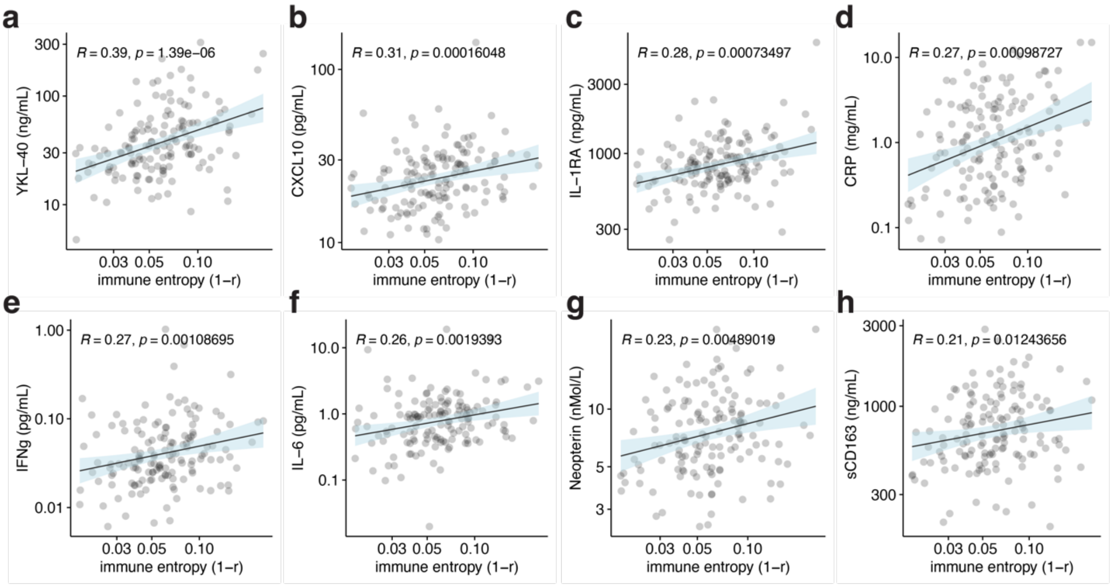


**Supplementary Figure 4:** Spearman correlations between immune entropy and concentrations of the following serum proteins: **(a)** YKL-40, **(b)** CXCL10, **(c)** IL-1RA, **(d)** CRP, **(e)** IFNg, **(f)** IL-6, **(g)** Neopterin, **(h)** sCD163.Correlations that were statistically significant after BH correction are reported. Axis ticks are log10 transformed.

**Supplementary Table 1**: Immune subset variables used in immune entropy calculation in VITAL and validation cohort

| VITAL cohort immune entropy variables (percentage of WBC) | Validation (ISA) cohort immune entropy variables (percentage of WBC) | RECOVAC cohort immune entropy variables (percentage of WBC) |
|---|---|---|
| granulocytes | granulocytes | |
| lymphocytes | lymphocytes | lymphocytes |
| CD3+ | CD3+ | CD3+ |
| CD4+ | CD4+ | CD4+ |
| CD4+ Tcm | CD4+ Tcm | CD4+ Tcm |
| CD4+ Tem | CD4+ Teff | CD4+ Tem |
| CD4+ Teff | CD4+ Tem | CD4+ Teff |
| CD4+ TrueNaive | CD4+ Tn | CD4+ TrueNaive |
| CD4+ Tscm | | CD4+ Tscm |
| CD4+ Treg | CD4+ Treg | CD4+ Treg |
| CD4+ Treg Tcm | CD4+ Treg Tcm | CD4+ Treg Tcm |
| CD4+ Treg Tem | CD4+ Treg Tem | CD4+ Treg Tem |
| CD4+ Treg Tn | CD4+ Treg Tn | CD4+ Treg Tn |
| CD4+ Treg CD38+ | | CD4+ Treg CD38+ |
| CD4+ Treg HLA-DR+ | | CD4+ Treg HLA-DR+ |
| CD4+ Treg CD95+ | | CD4+ Treg CD95+ |
| CD4+ CD38+ | CD4+ CD38+ | CD4+ CD38+ |
| CD4+ CD38+HLA-DR+ | CD4+ CD38+HLADR+ | CD4+ CD38+HLA-DR+ |
| CD4+ HLA-DR+ | CD4+HLA-DR+ | CD4+ HLA-DR+ |
| CD4+ CD95+ | | CD4+ CD95+ |
| CD4+ CXCR5+ | CD4+ CXCR5+ | CD4+ CXCR5+ |
| CD4+ CXCR5+CD38+ | | CD4+ CXCR5+CD38+ |
| CD4+ CXCR5+CD38-HLA-DR- | | CD4+ CXCR5+CD38-HLA-DR- |
| CD4+ CXCR5+CD95+ | | CD4+ CXCR5+CD95+ |
| CD4+ CXCR5+ Tcm | CD4+ CXCR5+ Tcm | CD4+ CXCR5+ Tcm |
| CD4+ CXCR5+ Tem | CD4+ CXCR5+ Tem | CD4+ CXCR5+ Tem |
| CD4+ CXCR5+ Tn | CD4+ CXCR5+ Tn | CD4+ CXCR5+ Tn |
| CD8+ | CD8+ | CD8+ |
| CD8+ Tcm | CD8+ Tcm | CD8+ Tcm |
| CD8+ Tem | CD8+ Teff | CD8+ Tem |
| CD8+ Teff | CD8+ Tem | CD8+ Teff |
| CD8+ TrueNaive | CD8+ Tn | CD8+ TrueNaive |
| CD8+ Tscm | | CD8+ Tscm |
| CD8+ CD38+ | CD8+ CD38+ | CD8+ CD38+ |
| CD8+ CD38+HLA-DR+ | CD8+ CD38+HLADR+ | CD8+ CD38+HLA-DR+ |
| CD8+ HLA-DR+ | CD8+HLA-DR+ | CD8+ HLA-DR+ |
| CD8+ CD95+ | | CD8+ CD95+ |
| CD8+ CXCR5+ | CD8+ CXCR5+ | CD8+ CXCR5+ |
| CD8+ CXCR5+CD95+ | | CD8+ CXCR5+CD95+ |
| CD8+ CXCR5+ Tcm | CD8+ CXCR5+ Tcm | CD8+ CXCR5+ Tcm |
| CD8+ CXCR5+ Tem | CD8+ CXCR5+ Tem | CD8+ CXCR5+ Tem |
| CD8+ CXCR5+ Tn | CD8+ CXCR5+ Tn | CD8+ CXCR5+ Tn |
| CD8+ CXCR5+ Teff | CD8+ CXCR5+ Teff | CD8+ CXCR5+ Teff |
| CD3- | CD3- | CD3- |
| CD56bright | CD56bright | |
| CD56bright CD38+ | CD56bright CD38+ | |
| CD56dim | CD56dim | |
| CD56dim CD38+ | CD56dim CD38+ | |
| CD56neg | CD56neg | |
| CD56neg HLA-DR+ | CD56neg HLA-DR+ | |
| CD19+ | CD19+ | CD19+ |
| B DN memory IgD-CD27- | B DN memory IgD-CD27- | B DN memory IgD-CD27- |
| B memory IgD+CD27+ | B memory IgD+CD27+ | B memory IgD+CD27+ |
| B naive IgD+CD27- | B naive IgD+CD27- | B naive IgD+CD27- |
| B switched memory IgD-CD27+ | B switched memory IgD-CD27+ | B switched memory IgD-CD27+ |
| monocytes | monocytes | monocytes |
| monocytes classical | monocytes classical | |
| monocytes intermediate | monocytes intermediate+nonclassical | |
| monocytes nonclassical | | |

**Supplementary Table 2**: Statistically significant differences in immune entropy between immunotypes. The significance was determined using Kruskal–Wallis tests. Post hoc tests were performed using Dunn's test with Benjamini-Hochberg method to adjust for multiple comparisons. *$P < 0.05$, **$P < 0.01$, ***$P < 0.001$, ****$P < 0.0001$

| variable | group1 | group2 | p.adj | p.adj.signif |
|---|---|---|---|---|
| immune entropy | immunotype 1 | immunotype 2 | 0.001942 | ** |
| immune entropy | immunotype 1 | immunotype 5 | 4.73E-05 | **** |
| immune entropy | immunotype 1 | immunotype 7 | 1.39E-06 | **** |
| immune entropy | immunotype 1 | immunotype 8 | 1.96E-10 | **** |
| immune entropy | immunotype 1 | immunotype 9 | 0.009396 | ** |
| immune entropy | immunotype 2 | immunotype 8 | 0.002278 | ** |
| immune entropy | immunotype 3 | immunotype 5 | 0.006693 | ** |
| immune entropy | immunotype 3 | immunotype 7 | 0.000514 | *** |
| immune entropy | immunotype 3 | immunotype 8 | 1.39E-06 | **** |
| immune entropy | immunotype 4 | immunotype 7 | 0.01111 | * |
| immune entropy | immunotype 4 | immunotype 8 | 0.000158 | *** |
| immune entropy | immunotype 5 | immunotype 6 | 0.009635 | ** |
| immune entropy | immunotype 5 | immunotype 8 | 0.042135 | * |
| immune entropy | immunotype 6 | immunotype 7 | 0.001027 | ** |
| immune entropy | immunotype 6 | immunotype 8 | 6.98E-06 | **** |
| immune entropy | immunotype 7 | immunotype 9 | 0.038277 | * |
| immune entropy | immunotype 8 | immunotype 9 | 0.000637 | *** |

**Supplementary Table 3**: Study characteristics of validation cohort (ISA), VITAL BNT162b2 cohort and kidney transplant patients (RECOVAC)

| Validation cohort (ISA) | Sex male (N=39) | Sex female (N=28) |
|---|---|---|
| age | 70 (67-73) | 71 (68-74) |
| CMV seropositivity | | |
| positive | 22 (56%) | 19 (68%) |
| negative | 17 (44%) | 9 (32%) |

| BNT162b2 VITAL cohort | Sex male (N=16) | Sex female (N=16) |
|---|---|---|
| age | 83 (82-86) | 82 (79-87) |
| CMV seropositivity | | |
| positive | 9 (56%) | 12 (75%) |
| negative | 7 (44%) | 4 (25%) |

| Kidney transplant patients (RECOVAC) | Sex male (N=30) | Sex female (N=29) |
|---|---|---|
| age | 56 (49-68) | 60 (53-69) |

Median (IQR); n (%)
